# Predictors of unsuccessful tuberculosis treatment outcome in Bhutan: A retrospective study using comprehensive national tuberculosis surveillance data

**DOI:** 10.1101/2025.08.05.25333000

**Authors:** Thinley Dorji, Karchung Tshering, Lila Adhikari, Thinley Jamtsho, Pavitra Bhujel, Pema Lhaden, Norelle L. Sherry, Chantel Lin, Justin T. Denholm, Sonam Wangchuk, Kristy Horan, Benjamin P. Howden, Patiyan Andersson

**Affiliations:** Kanglung Hospital, Trashigang, Ministry of Health, Bhutan; Department of Microbiology and Immunology at The Peter Doherty Institute for Infection and Immunity, University of Melbourne, Victoria, Australia; National Tuberculosis Reference Laboratory, Royal Centre for Disease Control, Ministry of Health, Serbithang, Thimphu; Royal Centre for Disease Control, Ministry of Health, Serbithang, Thimphu; Microbiological Diagnostic Unit Public Health Laboratory, Department of Microbiology and Immunology at The Peter Doherty Institute for Infection and Immunity, University of Melbourne, Victoria, Australia; Department of Infectious Diseases & Immunology, Austin Health, Heidelberg, Victoria, Australia; Royal Melbourne Hospital, Melbourne, Australia; Victorian Tuberculosis Program, Melbourne Health at the Peter Doherty Institute for Infection and Immunity, University of Melbourne, Melbourne, Australia; Department of Infectious Diseases at The Peter Doherty Institute for Infection and Immunity, University of Melbourne, Melbourne, Australia; Centre for Pathogen Genomics, University of Melbourne, Victoria, Australia

**Keywords:** Tuberculosis, Bhutan, treatment outcome, unsuccessful outcome

## Abstract

**Introduction:** Tuberculosis is a significant public health problem in Bhutan. Effective treatment is the key to TB elimination but information on treatment outcomes in Bhutan is limited. To improve TB control efforts in Bhutan, we assessed TB treatment outcomes and explored the factors that contribute to the unsuccessful treatment outcome using a comprehensive national TB dataset.

**Methods:** A retrospective cohort study was conducted to analyse the treatment outcome using national TB data for the period 2018-2021 provided by the National Tuberculosis Reference Laboratory, Royal Centre for Disease Control. Univariate and multiple logistic regression were performed to identify variables associated with an unsuccessful outcome.

**Results:** During the study period, 3,619 patients received TB treatment. Of these, 52.4% were pulmonary bacteriologically confirmed TB (PBC) and 27% were extra-pulmonary clinically diagnosed. Of the 3,330 patients with recorded treatment outcome, 96.2% had a successful treatment outcome (44.4% cured and 51.7% completed treatment) and 3.8% had an unsuccessful outcome (2.8% died; 0.4% lost to follow up; 0.7% treatment failure). Multiple logistic regression showed patients older than 60 years of age (aOR 4.3; 95% CI 2.11 – 10.1; p-value <0.001), diagnosed in 2021 (aOR 1.73; 95% CI 1.03 – 2.96; p-value 0.041), and PBC (aOR 2.33; 95% CI 1.49-3.8; p-value <0.001) were more likely to have an unsuccessful treatment outcome.

**Conclusion:** The proportion of successful TB treatment exceeded the global target rate of 90%, with a small proportion of unsuccessful TB treatment outcome. PBC and elderly patients require active follow-up to ensure that their treatment success increases. In-depth studies need to be conducted to understand circumstances leading to the treatment failures and death.

## INTRODUCTION

Tuberculosis (TB) remains a leading cause of death from a single infectious agent, especially in developing countries^1^. An estimated 23% of the global population have latent TB^2^ with approximately 5-10% of them expected to develop active disease over their lifetime^1^. Approximately, 10.6 million TB cases were recorded globally in 2022, with majority of them in Southeast Asia (46%), Africa (23%) and Western Pacific (18%)^1^.

TB is a public health problem in Bhutan with a prevalence rate of 190/100,000^3^. Globally, TB elimination is hampered by its long treatment duration, propensity to develop drug resistance and latency. Consequently, TB elimination is only possible through early detection and treatment with effective drugs to interrupt the transmission chain. The National Tuberculosis Control Program (NTCP) in Bhutan monitors the treatment outcome of all TB cases to ensure that patients receive timely treatment and are confirmed to be successfully cured. Treatment outcomes indicates the effectiveness of the TB control program^4^. Moreover, the availability of this data facilitates the comparison of TB program progress across different countries^5^ and monitor program targets and indicators. NTCP has adopted the WHO End-TB Strategy, which aims to reduce the TB incidence by 90% and TB deaths by 95% by 2035^3^.

Currently, there are few published studies assessing TB treatment outcome in Bhutan and those available are limited to single health centres^6,7^. Therefore, using comprehensive national TB notification data from 2018 to 2021, we aimed to: 1) evaluate the treatment outcome for all types of TB in Bhutan, and 2) identify the factors associated with unsuccessful treatment outcome.

## METHODS

### Setting

Health care services for the 20 districts in Bhutan are divided into three regions: Eastern, Western and Central region referral centres. Any presumptive TB patients undergo acid fast microscopy along with chest X-ray in the local health centres. Routine TB culture and phenotypic testing is managed by the National Tuberculosis Reference Laboratory (NTRL) at Royal Centre for Disease Control (RCDC), under the Ministry of Health. TB is a notifiable disease with mandatory reporting in the Tuberculosis Information Surveillance System (TBISS) before treatment commencement. This is a web-based surveillance system, managed by NTRL. Laboratory and epidemiological data for TB cases are recorded in this system by health workers from all TB centres. Cases are classified into the following categories: pulmonary bacteriologically confirmed (PBC) - a case of pulmonary TB with bacteriological confirmation through smear microscopy, Xpert MTB/RIF, or culture; extra-pulmonary bacteriologically confirmed (EPBC) - representing extrapulmonary tuberculosis with bacteriological confirmation; pulmonary clinically diagnosed (PCD) - referring to pulmonary TB clinically diagnosed in the absence of bacteriological confirmation; and extra-pulmonary clinically diagnosed (EPCD) - denoting extrapulmonary TB diagnosed clinically without bacteriological evidence. The TB cases are treated as per the National Tuberculosis Treatment Guidelines, which is aligned with WHO treatment recommendations^8^.

### Study design and population

This retrospective cohort study included all TB cases diagnosed and recorded in TBISS from 1 January 2018 to 31 December 2021. Patient information was de-identified by the data custodians at RCDC prior to being provided to the research team for analysis. Census data was obtained from the National Statistics Bureau (https://www.nsb.gov.bt/publications/annual-dzongkhag-statistics/) and World Bank database.

### Inclusion and exclusion criteria

We included all TB patients registered for treatment and recorded in the TBISS. In instances where patients were lost to follow up or treatment failed and re-treated in the same year, they were counted only once, with only the final treatment outcome recorded to avoid duplication. In the final analysis, patients whose treatment outcome was not known or recorded, were excluded.

### Case definition

A successful treatment outcome was defined as a patient with TB who was cured or had completed treatment (**Table 1**).^9^ An unsuccessful treatment outcome encompassed patients whose treatment failed, died, or were lost to follow-up. For PCD and EPCD TB, the treatment outcome was based on the clinical assessment and classification by a physician.

**Table 1.** Outcome definitions used in this paper. ^9^.

| Outcome | Definition |
| --- | --- |
| Cured | A case of PBC who completed treatment and confirmed to be bacteriologically negative in the last month of treatment or at least one previous occasion. |
| Treatment completed | A patient who completed the full course of treatment without evidence of failure, but without laboratory confirmation of smear in the last month of treatment or on completion of treatment. |
| Treatment failure | A patient who is smear positive or culture positive at five months of treatment or later. |
| Died | Patients with TB who died of any reason during or before treatment. |
| Lost to follow-up | Patients with TB who interrupted treatment for two or more consecutive months. |

### Variables used

The dependent variable was TB treatment outcome – dichotomously classified as successful or unsuccessful. The independent variables included for analysis were patient age, sex, region, year of diagnosis, type of TB, and treatment history. TB type was categorised into PBC, PCD, EPBC and EPCD.

### Epidemiological data analysis

All statistical analysis were performed using R 4.5.0. Frequencies and proportions were calculated for all the categorical variables. We used the Wilcoxon Rank-Sum test to compare age differences by sex, and the Cochran-Armitage test to assess trends in successful treatment outcomes by year. Non-parametric test such as Kruskal Wallis test was performed, followed by Dunn’s post hoc test with Bonferroni correction to test for differences in age by treatment outcome. Association of the dichotomous dependent variable with various indicators, were assessed in a multiple logistic regression model using glm. We used the stats::step() function in base R with the backward elimination method, which relies on the Akaike Information Criterion (AIC), to derive a parsimonious model. Backward elimination is preferred over forward and stepwise selection due to its ability to “asses joint predictive ability of variables”^10^. We chose the parsimonious models over complex model to reduce overfitting^10^. The model fit was tested with likelihood ratio test and Wald test, which showed no significant difference between full model and reduced model, supporting the parsimonious model. The strength of associations is presented as odds ratios with 95% confidence interval. The level of significance was set at p-value less than 0.05. In instances where multiple logistic regression could not be used due to small number of observations during crosstabulations, we used Firth’s penalized logistic regression^11^. Multicollinearity was tested using variance inflation factor (VIF) and all variables had VIF <1.5 and were retained in the analysis. Since DR-TB is associated with poor outcome^1^, we ran separate regression models. Initially, we ran a multiple logistic model without segregation. Then for latter part, we ran regression model separately for drug sensitive-TB, drug resistant-TB and PBC.

## Result

### Demographic characteristic

During the study period (2018-2021), a total of 3,619 patients received TB treatment. The median age of patients at the time of diagnosis was 28 years (range: 0-103 years), with the majority (64%) falling within the 18-39 years age group. The data also included 20 patients who were under six years of age, including five newborns treated for congenital TB. Females accounted for 52% (n=1,877) of TB cases. Male patients were significantly older than female patients, with a median age of 30 years versus 26 years, respectively (Wilcoxon rank-sum test, p < 0.001). Occupation groups were diverse, with students constituting the largest group (23.6%), followed by farmers (19.4%). In the study period, 33 (0.9%) health workers were also treated for TB.

### Geographical distribution of tuberculosis

The major portion of TB were notified from the Western region (82.7%), followed by the Central region (12.5%). The highest number of cases were diagnosed from Thimphu district (36.7%, n=1328), followed by Chhukha (12.2%, n=440), Sarpang district (8.6%, n=311), and Samtse (7%, n=253). The average incidence of TB during the study years was 117.7/100,000 population. After population standardisation, the highest incidence of TB was observed in Thimphu at 192.6/100,000 population, followed by Sarpang (162.9/100,00), Chhukha (158.4/100,000) and Monggar (147.5/100,000) (Fig 1).

**Figure 1.**
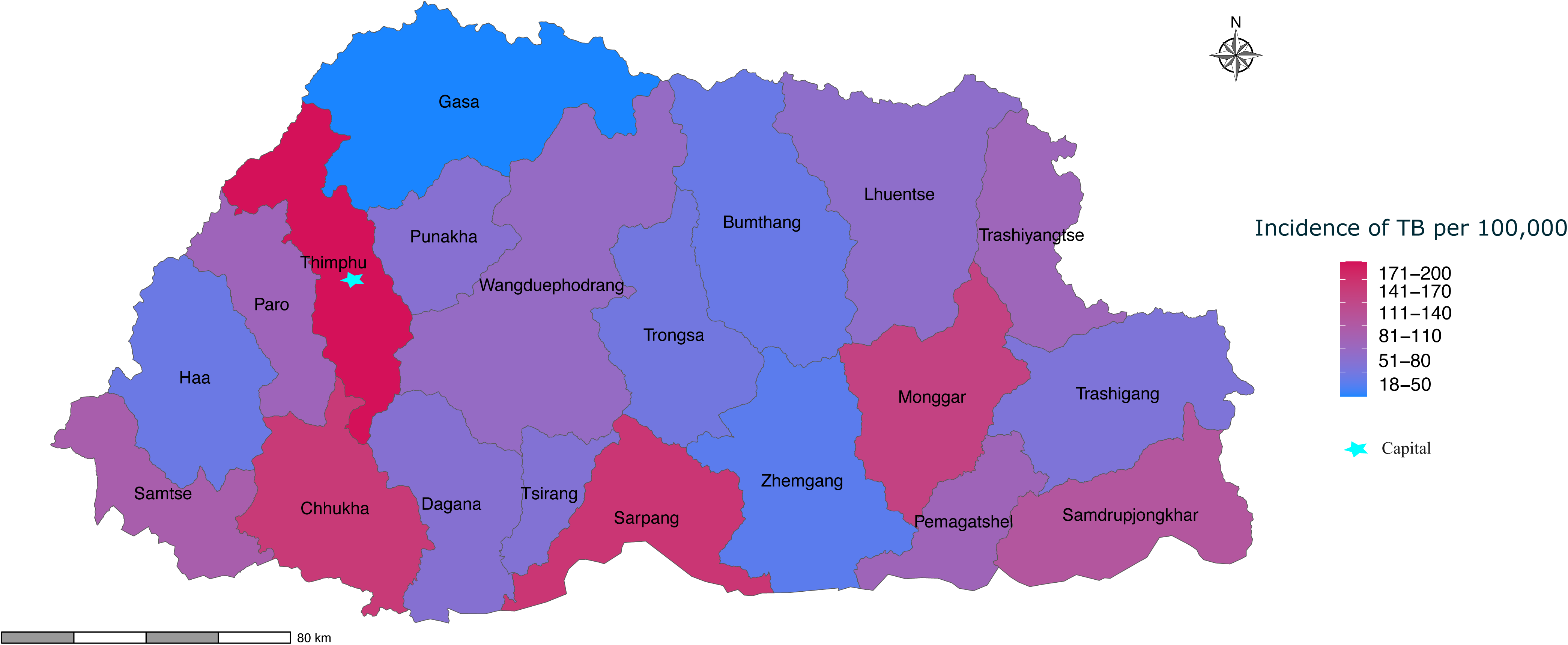
Map of Bhutan illustrating the district-wise distribution of tuberculosis (TB) cases from 2018 to 2021. The star symbol indicates the capital city. Incidence for each district is indicated with a blue (low) to red (high) gradient.

### Clinical characteristics

PBC was the most common TB (n=1,897, 52.4%) followed by EPCD (n=979, 27.1%) and EPBC (n=418, 11.6%). The majority of cases (n = 3,254; 89.9%) were diagnosed with TB for the first time, while 9.7% (n = 352) had a history of previous TB treatment. The treatment history for 13 cases were not known and this was all seen among those whose treatment outcome was not evaluated (Supplementary table 1).

### Treatment outcome based on type of cases

The dataset was divided into three groups based on whether the cases were drug-sensitive TB, any drug-resistant TB or clinically diagnosed/not tested TB (Fig 2). During the study period, 383 cases were resistant to at least one TB drug (rifampicin, isoniazid, ethambutol and streptomycin) based on Xpert MTB/RIF, line probe assay, or phenotypic drug susceptibility testing. A total of 1,906 were drug-sensitive TB while 1,330 cases were clinically diagnosed or not tested for resistance. We had 8% of data (n=289) where final treatment outcome was not evaluated (Supplementary table 1). This was predominantly seen during the year 2021, where 14% (n = 114) of patients did not have their treatment outcomes evaluated. 15 patients (0.5%) were re-treated in the same year (12 patients failed treatment and three lost to follow-up) and were included only once in the analysis. The successful treatment outcome was high for all three groups ranging from 97.8% for any drug-resistant TB to 95% for drug-sensitive TB. Deaths during the TB treatment accounted for the majority of unsuccessful treatment outcomes across all three groups (Fig 2).

**Figure 2.**
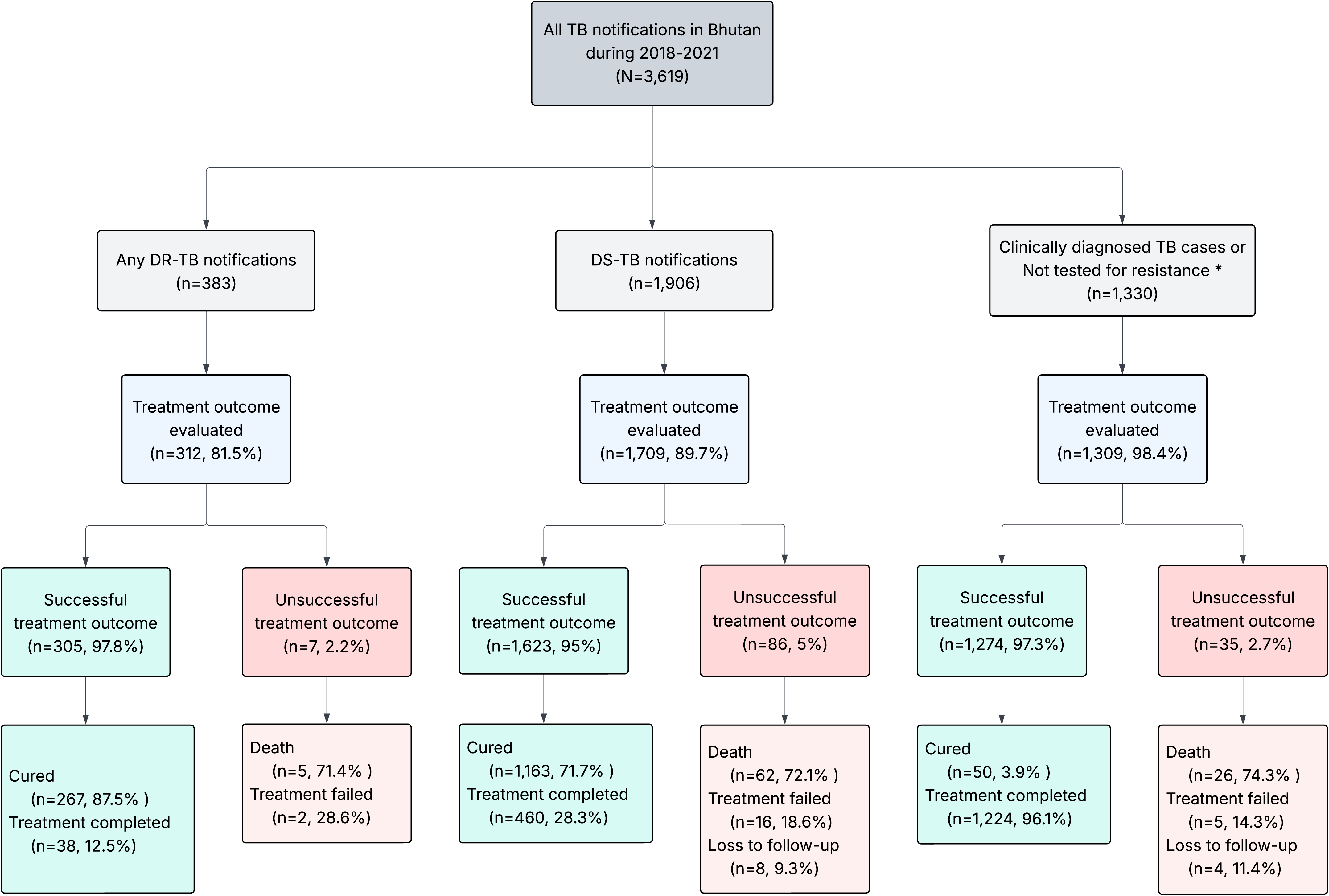
Flowchart depicting treatment outcome for various forms of TB. DR-TB (drug-resistant TB), DS-TB (drug-sensitive TB). *This also includes bacteriologically confirmed TB cases that were not tested for drug-resistance.

Overall, during the study period: 47.6% (n=1722) completed treatment, 40.9% (n=1480) were cured, 2.6% (n=93) died with TB, 0.6% (n=23) failed the treatment and 0.3% (n=12) were lost to follow-up during treatment. The proportions of different treatment outcome types varied among different districts (Supplementary figure 1). Treatment failures were seen mainly among patients from Bumthang, Dagana and Lhuentse, whereas most deaths were seen among patients from Trongsa, Trashigang and TrashiYangtse. The small number of cases in some districts prevented us from conducting further statistical analysis. We further explored the patient’s characteristics by the outcome types (Supplementary table 1). The median age of patients who died was 54 years (range: 36–69), which was significantly older (p < 0.05) than patients with other treatment outcomes, except lost to follow-up. Most patients who died were older than 60 years (46%, n=43), had PBC (68.8%, n=63) and were farmers (50.5%, n=47). The overall relationship between the year of diagnosis, region, type of case and site of infection with treatment outcome is summarised in Fig 3.

**Figure 3.**
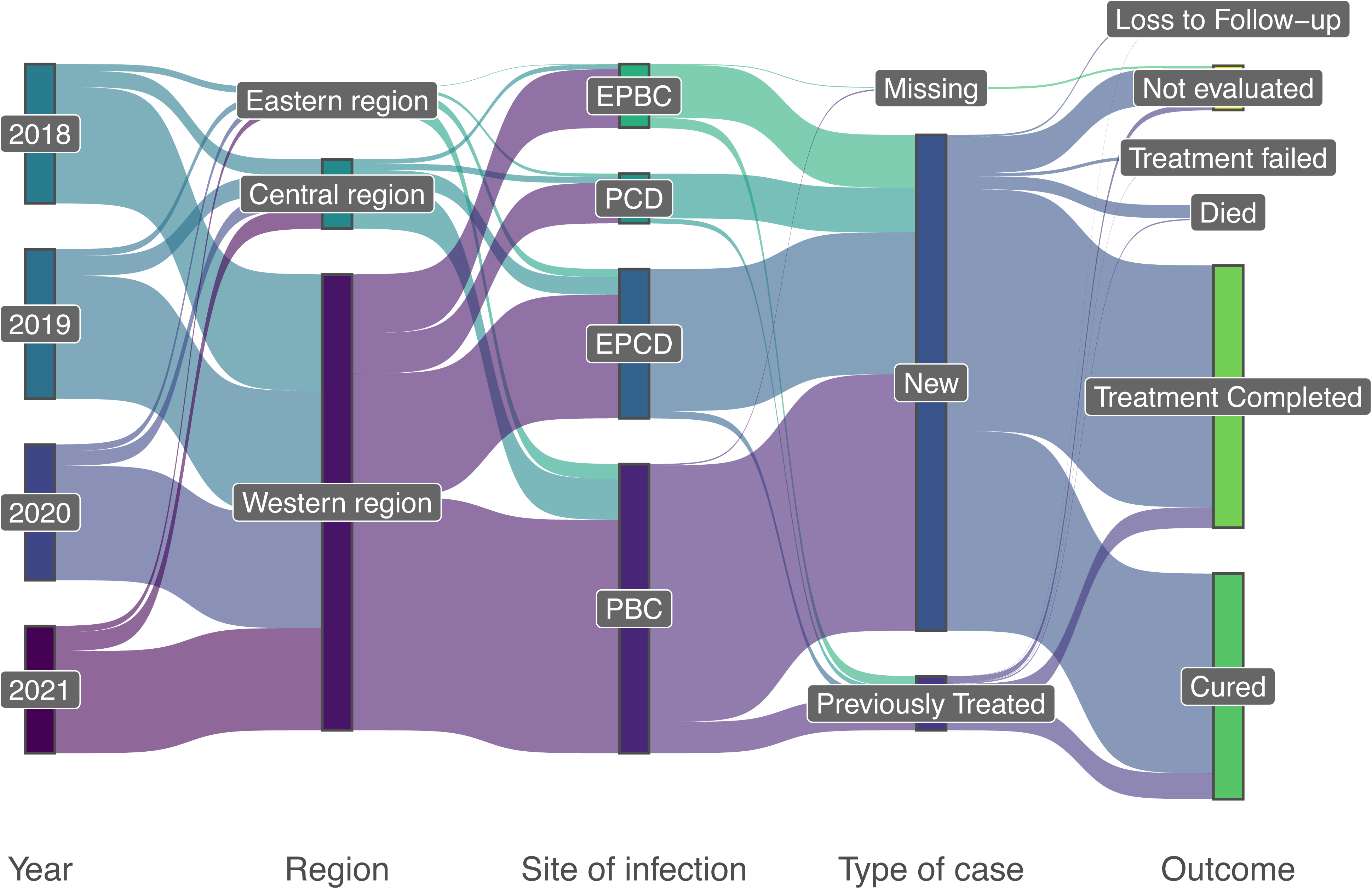
Sankey diagram summarising the TB treatment outcome by year of diagnosis, region, site of infection, and treatment history. PCD (pulmonary clinically diagnosed), PBC (pulmonary bacteriologically confirmed), EPCD (extra-pulmonary clinically diagnosed), EPBC (extra-pulmonary bacteriologically confirmed).

### Determinants of treatment outcome

Considering all available cases (including those with incomplete records for treatment outcome), the proportion of successful outcomes was 88.5%. After excluding cases without recorded treatment outcomes (n = 289, 8% of total cases), the overall treatment success rate increased to 96.2% (95% CI: 95.43% - 96.77%), with no significant variation over time (Cochran-Armitage test; p = 0.108).

Table 2 shows the results of univariate and multiple logistic regression assessing variables associated with unsuccessful outcome. Patients older than 60 years of age (aOR 4.3; 95% CI 2.11 – 10.1; p-value <0.001), diagnosed in 2021 (aOR 1.73; 95% CI 1.03 – 2.96; p-value 0.041), and PBC (aOR 2.33; 95% CI 1.49-3.8; p-value <0.001) were significantly more likely to have unsuccessful treatment outcome in the multiple logistic regression. Although male patients were 1.46 times more likely to have an unsuccessful outcome and those with EPBC were 79% less likely (OR 0.21) to experience unsuccessful treatment in the univariate analysis, these associations were not retained in the multivariable model.

**Table 2.**
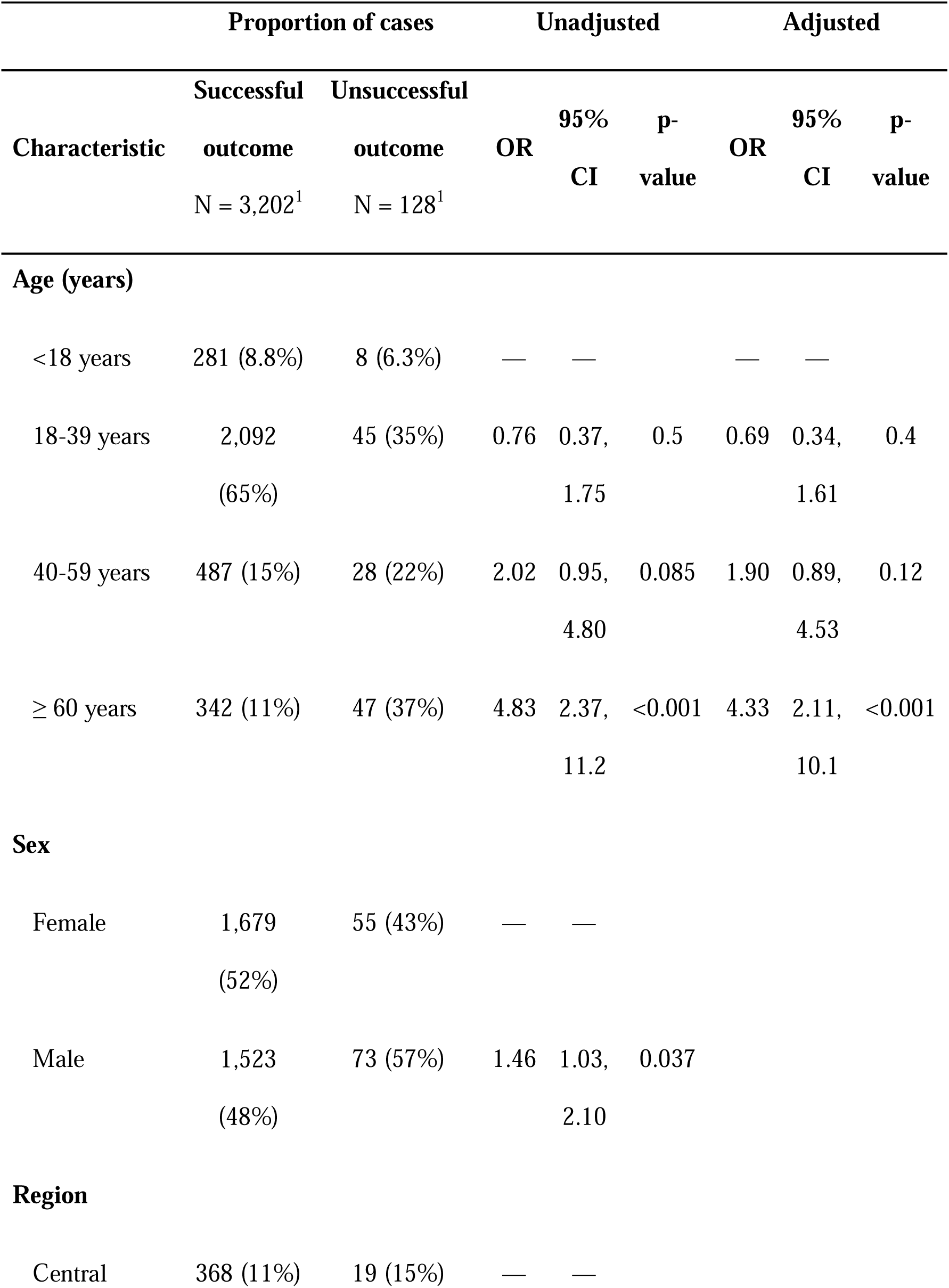
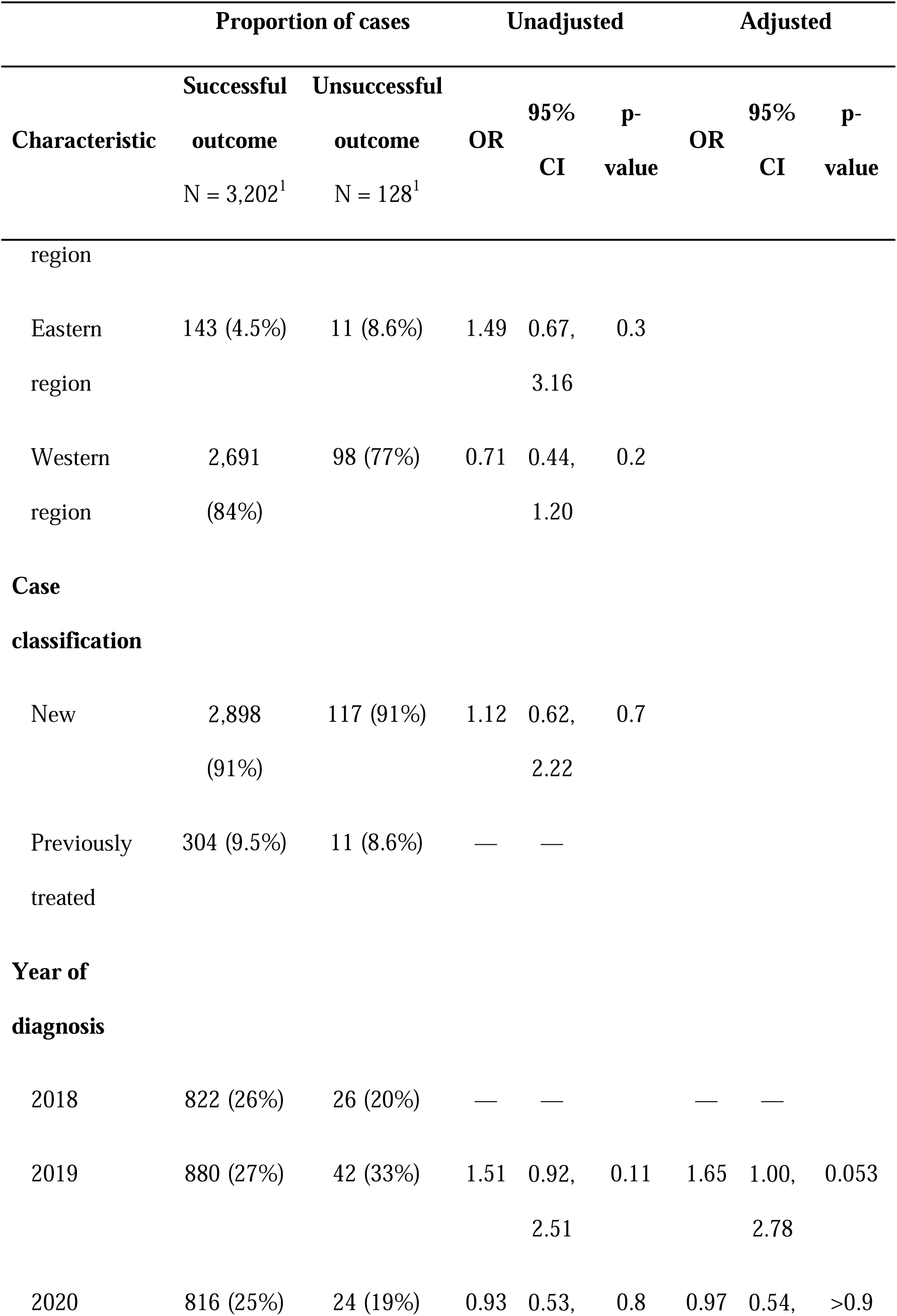
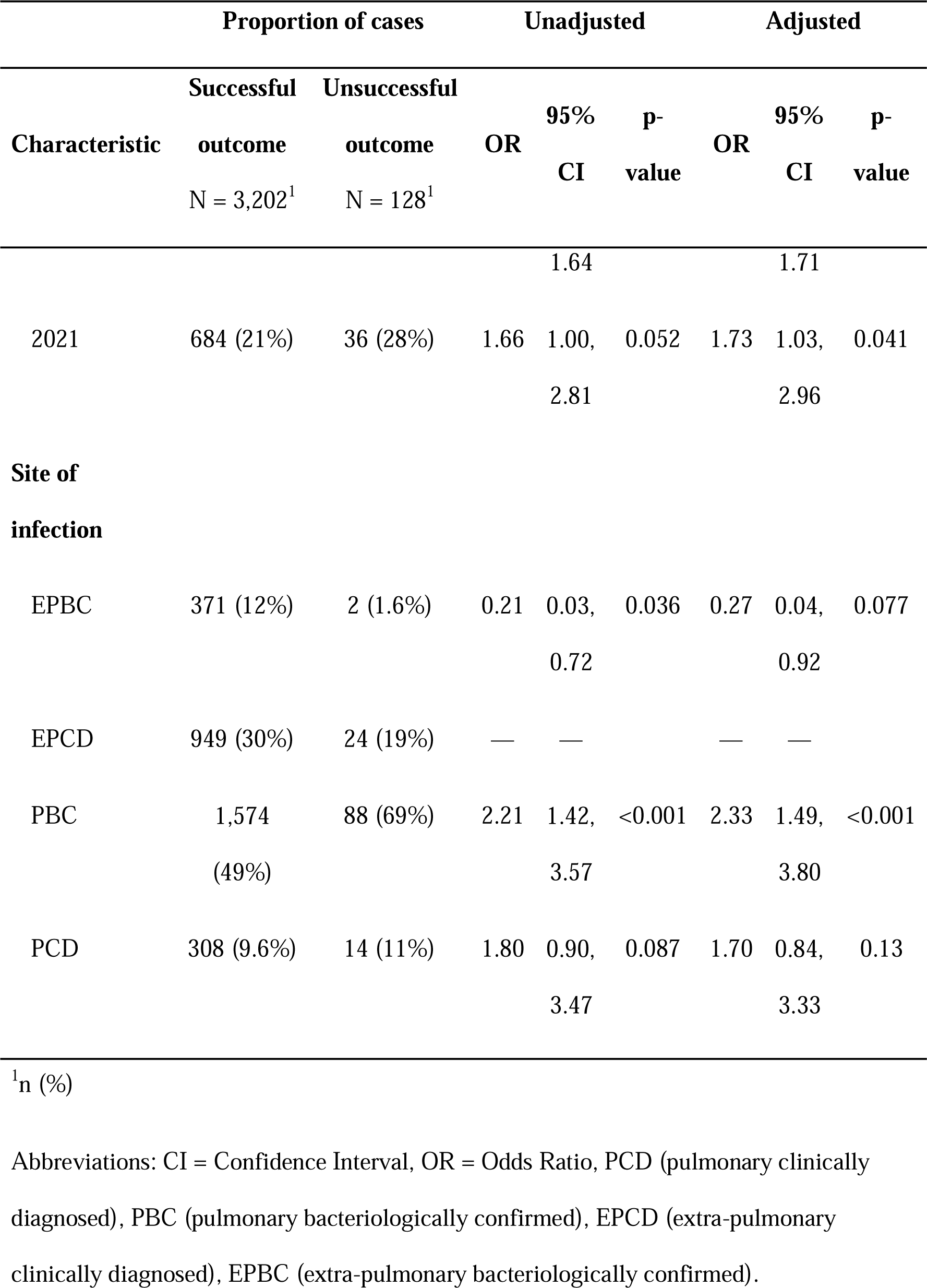
Determinants of successful treatment outcome in Bhutan.

We also investigated the treatment outcome specifically for PBC TB. The proportion of successful treatment among PBC TB was 94.7% (95% CI: 93.5% - 95.7%). Patients over the age of 60 (aOR 5.96, 95% CI 2.28-20.5, p-value 0.001) and TB cases diagnosed in 2021 (aOR 2.53, 95% CI 1.32-5.08, p-value 0.006) were more likely to have unsuccessful treatment outcome (supplementary table 2). Additionally, when we looked for factors associated with unsuccessful treatment outcome for drug-sensitive TB, we found that patients older than 60 years (aOR 7.8, 95% CI 2.72-33.0, p-value < 0.001) and patients aged 40-59 years (aOR 3.64, 95% CI 1.22, 15.7, p-value 0.04) (supplementary table 3) were more likely to have unsuccessful treatment outcome. For any drug-resistant TB, we applied Firth bias reduced logistic regression due to a fewer number of observations. However, no factors were found to be significantly associated with unsuccessful treatment outcome.

## Discussion

Using a comprehensive national TB data, this study demonstrated a high successful TB treatment outcome in Bhutan. Patients older than 60 years, diagnosed with TB in 2021, and with PBC were more likely to have unsuccessful TB treatment outcome.

Excluding the incomplete outcome data, the proportion of patients with unsuccessful treatment outcome in our dataset was low, with rate of successful outcome exceeding the global target of 90%^12^. This is consistent with previous studies in Bhutan assessing the treatment outcome at the level of health facilities, and among children, which showed low rate of unsuccessful outcome^6,7,13^. The low proportion of patients with unsuccessful treatment in this study is similar to studies from China^14^. This is likely due to the provision of free health care, and a regular follow-ups in Bhutan. Additionally, we also have high BCG vaccination coverage in Bhutan^15^, which is usually associated with good treatment outcome^16^. Although, we did not have data on BCG vaccination, we can postulate that most would have been vaccinated due to high coverage (99%) for more than two decades^15^. Finally, the low prevalence of HIV/AIDS in Bhutan^17^ likely contributes to the small proportion of unsuccessful TB treatment outcome^18^.

Despite the overall low proportion of unsuccessful treatment outcome, most were due to deaths (2.6%). While already lower than the case fatality rate reduction threshold proposed by WHO (6.5%) by 2025^19^, evidence-based approaches could further reduce these deaths.

Although we lacked information on timing of patients death during treatment, previous studies have shown that most deaths occur early in the treatment (< 2 months) and are often associated with PBC^20^. This highlights the importance of closely monitoring and follow-up of PBC cases during the intensive phase especially among the elderly patients. Delayed TB diagnosis and treatment are linked to higher mortality rates among TB patients^21^. The passive screening approach for TB used in Bhutan could affect timely diagnosis, and delays of 30 days has been reported previously^22^. Increased active TB screening for early diagnosis could have a considerable impact on reducing deaths - it would also likely have a positive effect on decreasing transmission. Non-adherence to treatment regimen has been shown to increase mortality rate from TB^23^. However, we do not have any information on adherence. Therefore, further in-depth verbal autopsy can be performed with regards to those who died to understand adherence and other risk factors. One proven method of improving drug adherence include electronic reminders as well as incentives^24^.

A high proportion of TB was observed among students, potentially a consequence of high contact with index cases and indoor transmission of the disease in classrooms and dormitories. Active case screening could be focussed among this cohort, particularly, in high burden districts of Bhutan. Furthermore, it is essential to increase awareness around TB symptoms and prevention in these settings, as knowledge about TB in Bhutan remains low^25^. We also observed health workers diagnosed with TB. Although this group accounts for fewer than 1%, their exposure remains a significant infection control concern, particularly due to the risks of transmitting the disease to vulnerable populations during service delivery. Despite evidence of regular screening leading to decrease in TB among the health workers^26^, there are currently no formal guidelines in place for screening of health workers for TB in Bhutan^3^.

Patients diagnosed and treated for TB in 2021 were more likely to have an unsuccessful outcome. This could be due to disruptions in TB management and care associated with lockdowns and reassignment of health workers to COVID-19 responses during the pandemic^27^. There was a deterioration in record keeping during the pandemic and TB case detection rate in Bhutan also dropped from 80% in 2019 to 67% in 2021^28^. Poor TB treatment outcomes during the pandemic were also described in other settings^29^ and an increase in TB deaths was observed globally^19^.

Consistent to our findings, PBC is associated with higher rates of unsuccessful outcomes compared to the other forms of TB^5,18^, due to high TB bacillary-load in the cavitary lesions of the lung^30^. Advanced age has been associated with unsuccessful TB treatment outcome^18,31^. This could be due to decrease in immunity as well as the development of co-morbidities. However, association with co-morbidities could not be assessed in this study as these data were not available.

Several factors favour Bhutan’s potential to achieve a significant reduction in TB burden. While Bhutan has a high prevalence of TB, the small population means that the absolute number of TB cases is low when compared to other high burden countries. Further, health care services are provided free of cost, and BCG vaccination is integrated within the expanded immunisation programme. As Bhutan strives towards achieving the End TB strategy, “patient-centered care”^32^ needs to be considered. Currently, PBC cases are admitted to isolation wards for few weeks to ensure interruption of transmission and patient drug compliance. However, recent evidence have shown no differences between health-worker administered directly observed treatment (DOT) and self-administered DOT^33^, and advocate for home-based isolations^34^ to prevent untoward mental stress, and stigmatization of the patient^35^. It is unlikely that home-based isolation would further exacerbate household transmission post-diagnosis, as the patient would be on treatment. An added benefit of home isolations is reduction of catastrophic costs (costs ≥ 20% of household annual income) as well as health system costs.

To our knowledge, this is the first study assessing the TB treatment outcome at the national level using a comprehensive national TB dataset. Nonetheless, there are some limitations. We lacked information on co-morbid illnesses, risky behaviours (eg. smoking and alcohol consumption), symptom duration, and drug adherence - factors that could influence the treatment outcomes. Routinely collected surveillance data will have varying degrees of completeness. Some important indicators known to influence treatment outcome such as weight^36^, were not recorded in the online reporting system for the majority of cases and was dropped from the analysis. Approximately 8% of the outcome data were not updated in the system by the health workers and hence had to be excluded during the final analysis.

Improved completeness and accuracy of the record keeping would support real-time monitoring and enhance management of TB patients. Further, as disease incidence decreases, more variables could be incorporated in the surveillance system to inform the design and strategies to further improve TB treatment adherence. In our dataset most cases were PBC, but for a considerable proportion the treatment outcome was listed as treatment completed rather than cured. Although, both cure and treatment completed show good prognosis, it is best practice for PBC to be bacteriologically declared negative after completion of treatment^9^.Therefore, testing protocols and subsequent documentation of bacteriologic confirmation of cure needs to be enhanced.

## Conclusion

For the first time, we present the treatment outcome rates using comprehensive national data for the years 2018-2021 in Bhutan. The findings from this study can be used to formulate support services and improve monitoring of the patients who are at high risk of treatment failure. Despite Bhutan’s high treatment success rate, there remains potential to further improve treatment outcome through decreasing TB mortality. Additionally, documentation in the online surveillance system could be strengthened. Our findings were generated using the national data and therefore can be used for inter-country comparison across the region with similar health system.

## Contributors

TD, KH, BPH and PA conceived the project, data analysis and drafting manuscript. KT, LA, TJ, PB, PL and SW were involved in data collection, preparation and finalisation of manuscript. CL, JD and NS were involved in analysis and finalisation of the manuscript. All authors read and approved the final manuscript.

## Acknowledgement

We are grateful to the Ministry of Health, Bhutan for granting us the permission to carry out this study and to Royal Centre for Disease Control for giving us access to the data.

## Ethics approval

The study was approved by the Research and Ethics Board of Health, Ministry of Health, Bhutan (REBH/Approval/2022/027) and the University of Melbourne (Approval reference no. 2023-26130-41858-3). Waiver for informed consent was applied as this was a retrospective study.

## Data availability statement

Data requests can be made to the corresponding author, with sharing subject to the discretion of the Royal Centre for Disease Control, Bhutan, as the data custodians.

