## Supplementary figures and images for "Predictors of unsuccessful tuberculosis treatment outcome in Bhutan: A retrospective study using comprehensive national tuberculosis surveillance data"

### Supplementary figure 1

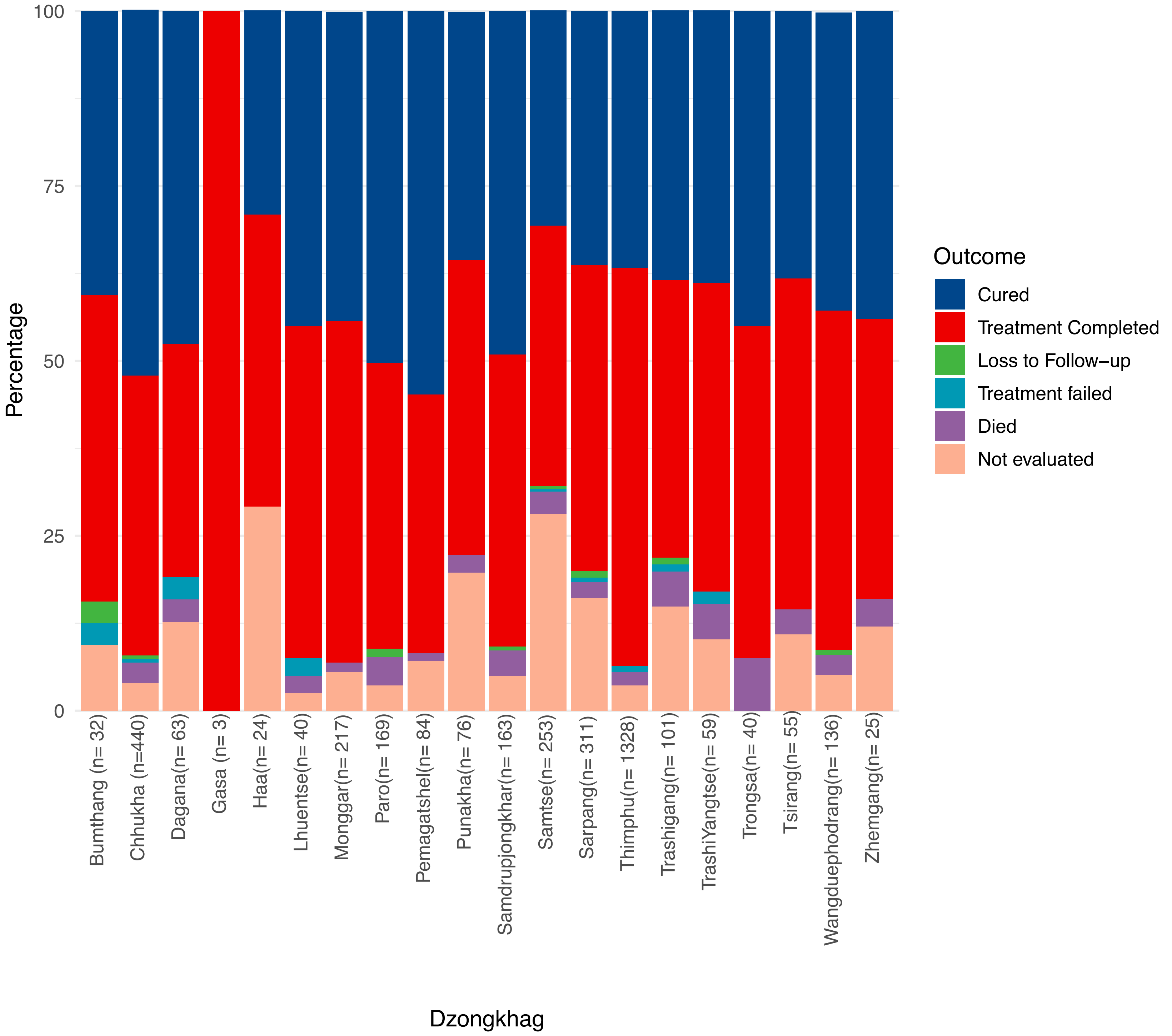
