## Supplementary table 1 for "Predictors of unsuccessful tuberculosis treatment outcome in Bhutan: A retrospective study using comprehensive national tuberculosis surveillance data"

Supplementary table 1. Sociodemographic and clinical characteristics by treatment outcome types.

| **Characteristic** | **Cured**  N = 1,480^1^ | **Treatment Completed**  N = 1,722^1^ | **Treatment failed**  N = 23^1^ | **Died**  N = 93^1^ | **Loss to Follow-up**  N = 12^1^ | **Not evaluated**  N = 289^1^ | **Overall**  N = 3,619^1^ |
| --- | --- | --- | --- | --- | --- | --- | --- |
| **Age (median)** | 27 (22, 39) | 28 (21, 41) | 29 (23, 44) | 54 (36, 69) | 37 (22, 43) | 27 (21, 44) | 28 (22, 42) |
| **Age** |  |  |  |  |  |  |  |
| <18 years | 101 (6.8%) | 180 (10%) | 4 (17%) | 3 (3%) | 1 (8.3%) | 30 (10%) | 319 (8.8%) |
| 18-39 years | 1,011 (68%) | 1,081 (63%) | 13 (57%) | 25 (27%) | 7 (58%) | 166 (57%) | 2,303 (64%) |
| 40-59 years | 219 (15%) | 268 (16%) | 3 (13%) | 22 (24%) | 3 (25%) | 64 (22%) | 579 (16%) |
| ≥ 60 years | 149 (10%) | 193 (11%) | 3 (13%) | 43 (46%) | 1 (8.3%) | 29 (10%) | 418 (12%) |
| **Gender** |  |  |  |  |  |  |  |
| Female | 748 (51%) | 931 (54%) | 12 (52%) | 40 (43%) | 3 (25%) | 143 (49%) | 1,877 (52%) |
| Male | 732 (49%) | 791 (46%) | 11 (48%) | 53 (57%) | 9 (75%) | 146 (51%) | 1,742 (48%) |
| **Treatment history** |  |  |  |  |  |  |  |
| Missing | 0 (0%) | 0 (0%) | 0 (0%) | 0 (0%) | 0 (0%) | 13 (4.5%) | 13 (0.4%) |
| New | 1,311 (89%) | 1,587 (92%) | 21 (91%) | 85 (91%) | 11 (92%) | 239 (83%) | 3,254 (90%) |
| Previously Treated | 169 (11%) | 135 (7.8%) | 2 (8.7%) | 8 (8.6%) | 1 (8.3%) | 37 (13%) | 352 (9.7%) |
| **Region** |  |  |  |  |  |  |  |
| Central region | 175 (12%) | 193 (11%) | 4 (17%) | 12 (13%) | 3 (25%) | 67 (23%) | 454 (13%) |
| Eastern region | 70 (4.7%) | 73 (4.2%) | 3 (13%) | 6 (6.5%) | 2 (17%) | 19 (6.6%) | 173 (4.8%) |
| Western region | 1,235 (83%) | 1,456 (85%) | 16 (70%) | 75 (81%) | 7 (58%) | 203 (70%) | 2,992 (83%) |
| **Year of diagnosis** |  |  |  |  |  |  |  |
| 2018 | 368 (25%) | 454 (26%) | 6 (26%) | 19 (20%) | 1 (8.3%) | 66 (23%) | 914 (25%) |
| 2019 | 430 (29%) | 450 (26%) | 9 (39%) | 28 (30%) | 5 (42%) | 59 (20%) | 981 (27%) |
| 2020 | 379 (26%) | 437 (25%) | 3 (13%) | 20 (22%) | 1 (8.3%) | 50 (17%) | 890 (25%) |
| 2021 | 303 (20%) | 381 (22%) | 5 (22%) | 26 (28%) | 5 (42%) | 114 (39%) | 834 (23%) |
| **Occupation** |  |  |  |  |  |  |  |
| Armed forces | 23 (1.6%) | 33 (1.9%) | 1 (4.3%) | 0 (0%) | 0 (0%) | 5 (1.7%) | 62 (1.7%) |
| Civil Servant | 76 (5.1%) | 119 (6.9%) | 0 (0%) | 3 (3.2%) | 0 (0%) | 11 (3.8%) | 209 (5.8%) |
| Corporate employee | 33 (2.2%) | 44 (2.6%) | 0 (0%) | 0 (0%) | 0 (0%) | 7 (2.4%) | 84 (2.3%) |
| Dependent | 15 (1.0%) | 24 (1.4%) | 0 (0%) | 1 (1.1%) | 0 (0%) | 1 (0.3%) | 41 (1.1%) |
| Driver | 25 (1.7%) | 31 (1.8%) | 1 (4.3%) | 1 (1.1%) | 1 (8.3%) | 3 (1.0%) | 62 (1.7%) |
| Farmer | 257 (17%) | 312 (18%) | 6 (26%) | 47 (51%) | 3 (25%) | 78 (27%) | 703 (19%) |
| Housewife | 152 (10%) | 190 (11%) | 3 (13%) | 7 (7.5%) | 0 (0%) | 30 (10%) | 382 (11%) |
| Labour | 3 (0.2%) | 1 (<0.1%) | 0 (0%) | 0 (0%) | 2 (17%) | 3 (1.0%) | 9 (0.2%) |
| Minors | 0 (0%) | 26 (1.5%) | 0 (0%) | 2 (2.2%) | 0 (0%) | 0 (0%) | 28 (0.8%) |
| Monk/Nun | 19 (1.3%) | 31 (1.8%) | 0 (0%) | 1 (1.1%) | 1 (8.3%) | 4 (1.4%) | 56 (1.5%) |
| Others | 272 (18%) | 223 (13%) | 3 (13%) | 5 (5.4%) | 1 (8.3%) | 45 (16%) | 549 (15%) |
| Prisoner | 11 (0.7%) | 20 (1.2%) | 0 (0%) | 0 (0%) | 0 (0%) | 1 (0.3%) | 32 (0.9%) |
| Private /Business | 196 (13%) | 205 (12%) | 1 (4.3%) | 2 (2.2%) | 2 (17%) | 26 (9.0%) | 432 (12%) |
| Retiree | 53 (3.6%) | 40 (2.3%) | 2 (8.7%) | 17 (18%) | 0 (0%) | 4 (1.4%) | 116 (3.2%) |
| Student/Trainee | 345 (23%) | 423 (25%) | 6 (26%) | 7 (7.5%) | 2 (17%) | 71 (25%) | 854 (24%) |
| **Site of infection** |  |  |  |  |  |  |  |
| EPBC | 0 (0%) | 371 (22%) | 1 (4.3%) | 0 (0%) | 1 (8.3%) | 45 (16%) | 418 (12%) |
| EPCD | 0 (0%) | 949 (55%) | 4 (17%) | 18 (19%) | 2 (17%) | 6 (2.1%) | 979 (27%) |
| PBC | 1,479 (100%) | 95 (5.5%) | 16 (70%) | 64 (69%) | 8 (67%) | 235 (81%) | 1,897 (52%) |
| PCD | 1 (<0.1%) | 307 (18%) | 2 (8.7%) | 11 (12%) | 1 (8.3%) | 3 (1.0%) | 325 (9.0%) |
| ^1^Median (Q1, Q3); n (%) | | | | | | | |
