## Supplementary table 2 for "Predictors of unsuccessful tuberculosis treatment outcome in Bhutan: A retrospective study using comprehensive national tuberculosis surveillance data"

Supplementary table 1. Determinants of un-successful treatment outcome in PBC

|  | **Proportion of cases** | | **Unadjusted** | | | **Adjusted** | | |
| --- | --- | --- | --- | --- | --- | --- | --- | --- |
| **Characteristic** | **Successful outcome**  N = 1,574^1^ | **Unsuccessful outcome**  N = 88^1^ | **OR** | **95% CI** | **p-value** | **OR** | **95% CI** | **p-value** |
| **Age (years)** |  |  |  |  |  |  |  |  |
| <18 years | 111 (7.1%) | 4 (4.5%) | — | — |  | — | — |  |
| 18-39 years | 1,070 (68%) | 31 (35%) | 0.80 | 0.31, 2.74 | 0.7 | 0.81 | 0.31, 2.75 | 0.7 |
| 40-59 years | 229 (15%) | 19 (22%) | 2.30 | 0.84, 8.08 | 0.14 | 2.39 | 0.87, 8.42 | 0.12 |
| ≥ 60 years | 164 (10%) | 34 (39%) | 5.75 | 2.22, 19.7 | 0.001 | 5.96 | 2.28, 20.5 | 0.001 |
| **Gender** |  |  |  |  |  |  |  |  |
| Female | 794 (50%) | 38 (43%) | — | — |  |  |  |  |
| Male | 780 (50%) | 50 (57%) | 1.34 | 0.87, 2.08 | 0.2 |  |  |  |
| **Region** |  |  |  |  |  |  |  |  |
| Central region | 195 (12%) | 14 (16%) | — | — |  |  |  |  |
| Eastern region | 73 (4.6%) | 6 (6.8%) | 1.14 | 0.39, 2.97 | 0.8 |  |  |  |
| Western region | 1,306 (83%) | 68 (77%) | 0.73 | 0.41, 1.37 | 0.3 |  |  |  |
| **Case classification** |  |  |  |  |  |  |  |  |
| New | 1,391 (88%) | 82 (93%) | 1.80 | 0.84, 4.67 | 0.2 | 2.11 | 0.96, 5.56 | 0.091 |
| Previously treated | 183 (12%) | 6 (6.8%) | — | — |  | — | — |  |
| **Year of diagnosis** |  |  |  |  |  |  |  |  |
| 2018 | 397 (25%) | 14 (16%) | — | — |  | — | — |  |
| 2019 | 451 (29%) | 24 (27%) | 1.51 | 0.78, 3.03 | 0.2 | 1.70 | 0.87, 3.47 | 0.13 |
| 2020 | 395 (25%) | 20 (23%) | 1.44 | 0.72, 2.94 | 0.3 | 1.48 | 0.73, 3.08 | 0.3 |
| 2021 | 331 (21%) | 30 (34%) | 2.57 | 1.37, 5.07 | 0.004 | 2.53 | 1.32, 5.08 | 0.006 |
| ^1^n (%) | | | | | | | | |
| Abbreviations: CI = Confidence Interval, OR = Odds Ratio | | | | | | | | |
