## Supplementary table 3 for "Predictors of unsuccessful tuberculosis treatment outcome in Bhutan: A retrospective study using comprehensive national tuberculosis surveillance data"

Supplementary table 3. Determinants of un-successful treatment outcome among DS-TB in Bhutan.

|  | **Proportion of cases** | | **Unadjusted** | | | **Adjusted** | | |
| --- | --- | --- | --- | --- | --- | --- | --- | --- |
| **Characteristic** | **Successful outcome**  N = 1,623^1^ | **Unsuccessful outcome**  N = 86^1^ | **OR** | **95% CI** | **p-value** | **OR** | **95% CI** | **p-value** |
| **Age (years)** |  |  |  |  |  |  |  |  |
| <18 years | 117 (7.2%) | 3 (3.5%) | — | — |  | — | — |  |
| 18-39 years | 1,127 (69%) | 30 (35%) | 1.04 | 0.36, 4.38 | >0.9 | 1.04 | 0.36, 4.38 | >0.9 |
| 40-59 years | 214 (13%) | 20 (23%) | 3.64 | 1.22, 15.7 | 0.040 | 3.64 | 1.22, 15.7 | 0.040 |
| ≥ 60 years | 165 (10%) | 33 (38%) | 7.80 | 2.72, 33.0 | <0.001 | 7.80 | 2.72, 33.0 | <0.001 |
| **Gender** |  |  |  |  |  |  |  |  |
| Female | 863 (53%) | 36 (42%) | — | — |  |  |  |  |
| Male | 760 (47%) | 50 (58%) | 1.58 | 1.02, 2.46 | 0.042 |  |  |  |
| **Region** |  |  |  |  |  |  |  |  |
| Central region | 175 (11%) | 12 (14%) | — | — |  |  |  |  |
| Eastern region | 67 (4.1%) | 6 (7.0%) | 1.31 | 0.44, 3.51 | 0.6 |  |  |  |
| Western region | 1,381 (85%) | 68 (79%) | 0.72 | 0.40, 1.42 | 0.3 |  |  |  |
| **Case classification** |  |  |  |  |  |  |  |  |
| New | 1,449 (89%) | 78 (91%) | 1.17 | 0.59, 2.67 | 0.7 |  |  |  |
| Previously treated | 174 (11%) | 8 (9.3%) | — | — |  |  |  |  |
| **Year of diagnosis** |  |  |  |  |  |  |  |  |
| 2018 | 382 (24%) | 14 (16%) | — | — |  |  |  |  |
| 2019 | 413 (25%) | 22 (26%) | 1.45 | 0.74, 2.95 | 0.3 |  |  |  |
| 2020 | 443 (27%) | 22 (26%) | 1.36 | 0.69, 2.75 | 0.4 |  |  |  |
| 2021 | 385 (24%) | 28 (33%) | 1.98 | 1.05, 3.93 | 0.041 |  |  |  |
| ^1^n (%) | | | | | | | | |
| Abbreviations: CI = Confidence Interval, OR = Odds Ratio | | | | | | | | |
