## Supplementary table 4 for "Predictors of unsuccessful tuberculosis treatment outcome in Bhutan: A retrospective study using comprehensive national tuberculosis surveillance data"

Supplementary table 4. Determinants of successful treatment outcome among any drug-resistant TB in Bhutan.

|  | **Proportion of cases** | | **Unadjusted** | | | **Adjusted** | | |
| --- | --- | --- | --- | --- | --- | --- | --- | --- |
| **Characteristic** | **Successful outcome**  N = 305^1^ | **Unsuccessful outcome**  N = 7^1^ | **OR** | **95% CI** | **p-value** | **OR** | **95% CI** | **p-value** |
| **Age (years)** |  |  |  |  |  |  |  |  |
| <18 years | 17 (5.6%) | 1 (14%) | — | — |  |  |  |  |
| 18-39 years | 225 (74%) | 4 (57%) | 0.233 | 0.04, 2.41 | 0.186 | 0.339 | 0.05, 3.70 | 0.323 |
| 40-59 years | 46 (15%) | 1 (14%) | 0.376 | 0.029, 4.87 | 0.418 | 0.791 | 0.05, 12.04 | 0.84 |
| ≥ 60 years | 17 (5.6%) | 1 (14%) | 1.00 | 0.076, 13.2 | 1 | 2.29 | 0.12,46.05 | 0.561 |
| **Sex** |  |  |  |  |  |  |  |  |
| Female | 156 (51%) | 5 (71%) | — | — |  |  |  |  |
| Male | 149 (49%) | 2 (29%) | 0.476 | 0.085, 2.01 | 0.319 | 0.346 | 0.047, 1.779 | 0.212 |
| **Region** |  |  |  |  |  |  |  |  |
| Central region | 35 (11%) | 2 (29%) | — | — |  |  |  |  |
| Eastern region | 13 (4.3%) | 0 (0%) | 0.526 | 0.004, 7.03 | 0.666 | 0.643 | 0.004, 10.86 | 0.782 |
| Western region | 257 (84%) | 5 (71%) | 0.303 | 0.07, 1.74 | 0.161 | 0.251 | 0.053, 1.485 | 0.117 |
| **Case classification** |  |  |  |  |  |  |  |  |
| New | 258 (85%) | 7 (100%) | 2.76 | 0.326, 360. | 0.424 | 1.96 | 0.215, 257.38 | 0.62 |
| Previously Treated | 47 (15%) | 0 (0%) |  |  |  |  |  |  |
| **Year of diagnosis** |  |  |  |  |  |  |  |  |
| 2018 | 85 (28%) | 1 (14%) | — | — |  |  |  |  |
| 2019 | 104 (34%) | 3 (43%) | 1.91 | 0.307, 20.0 | 0.498 | 2 | 0.316, 21.15 | 0.468 |
| 2020 | 62 (20%) | 0 (0%) | 0.456 | 0.003, 8.69 | 0.613 | 0.443 | 0.003, 8.17 | 0.597 |
| 2021 | 54 (18%) | 3 (43%) | 3.66 | 0.584, 38.5 | 0.168 | 3.4 | 0.539, 35.25 | 0.195 |
| ^1^n (%), CI = Confidence Interval, OR = Odds Ratio | | | | | | | | |
| Due to small sample size and rare events, we used Firth’s penalized logistic regression using the r-package logistf. | | | | | | | | |
